# Effective Screening of SARS-CoV-2 Neutralizing Antibodies in Patient Serum using Lentivirus Particles Pseudotyped with SARS-CoV-2 Spike Glycoprotein

**DOI:** 10.1101/2020.05.21.20108951

**Authors:** Ritesh Tandon, Dipanwita Mitra, Poonam Sharma, Stephen S. Stray, John T. Bates, Gailen D. Marshall

**Author notes:** Ritesh Tandon **Email:**.

## Abstract

Pseuodotyped particles have significant importance and use in virology as tools for studying the biology of highly pathogenic viruses in a lower biosafety environment. The biological, chemical, and serological studies of the recently emerged SARS-CoV-2 will be greatly aided by the development and optimization of a suitable pseudotyping system. Here, we pseudotyped the SARS-CoV-2 Spike glycoprotein (SPG) on a retroviral (MMLV) as well as a third generation lentiviral (pLV) vector and tested the transduction efficiency in several mammalian cell lines expressing SARS-CoV-2 receptor hACE2. While MMLV pseudotyped the vesicular stomatitis virus G glycoprotein (VSV-G) efficiently, it could not pseudotype SPG. In contrast, pLV pseudotyped both glycoproteins efficiently; however, much higher titers of pLV-G particles were produced. Among all the tested mammalian cells, 293Ts expressing hACE2 were most efficiently transduced using the pLV-S system. The pLV-S particles were efficiently neutralized by diluted serum (>:640) from a recently recovered COVID-19 patient who showed high SARS-CoV-2 specific IgM and IgG levels. In summary, pLV-S pseudotyped virus provides a valid screening tool for the presence of anti SARS-CoV-2 specific neutralizing antibodies in convalescent patient serum.

**Significance Statement:** SARS-CoV-2 has emerged as one of the biggest threats in the history of humankind and is comparable to medieval plague, 1918 Spanish Flu, as well as world wars. Investigations into the biology of SARS-CoV-2 are partially hindered by the highly transmissible and pathogenic nature of this virus, which requires biosafety level 3 containment in a laboratory for investigation. The study here describes a pseudotyping system which mimics the surface properties of SARS-CoV-2 and can be used in lower biosafety level laboratory for the purpose of vaccine studies, drug inhibition studies, and serological screening to determine the status of herd immunity.

## Introduction

The recent emergence of Severe Acute Respiratory Syndrome Coronavirus (SARS-CoV-2) in Wuhan, China in late 2019 and its subsequent spread to the rest of the world has created a pandemic situation unprecedented in modern history (1-4). While initial challenges included diagnosis and proper containment of the infection, contemporary efforts are directed towards the measurement of antiviral total and neutralizing antibodies in recovered and symptomatic patients as well as in non-symptomatic vulnerable population (5-7). Studying the development of herd immunity, effectiveness of various upcoming vaccine candidates as well as establishing social parameters for re-opening of the world economy all depend on our ability to accurately measure neutralizing antibodies and establish the kinetics of their persistence in serum. Convalescent plasma therapy has shown early therapeutic successes in severe COVID-19 patients (8). Thus, the determination of neutralizing antibody titers in convalescent serum should provide significant assistance in clinical decision making regarding transfusion for therapeutic and possibly prophylactic indications.

Due to the highly transmissible and pathogenic nature of SARS-CoV-2, handling of live virus requires a biosafety level 3 (BSL3) containment (9). Thus, only facilities equipped with BSL-3 can safely study neutralizing responses using live virus. In order to extend this capability to other BSL-2 laboratories that are more widely available, a safer way is needed to measure both total antiviral antibody levels based on ELISAs and neutralizing antibody responses in a practical, reproducible surrogate assay that effectively replaces the need for the live SARS-CoV-2. To address this issue, we report the development of a high titer pseudotype virus that can be easily produced and successfully employed to screen patient serum for neutralizing antibodies in a lower biosafety level laboratory. Since the backbone of this virus consists of a non-replicating lentivirus, it poses no risk of infection to the personnel involved and the technology can readily be scaled up to the levels where it can be employed for screening of large sample numbers. These pseudotyped particles (pLV-S) can also be used for immunological, pharmacological and biochemical studies as it represents SPG in its native conformation.

## Results

### SARS-CoV-2 Spike protein can be efficiently pseudotyped on a lentiviral vector

We used a traditional retroviral pseudotyping system based on Murine Molony Leukemic Virus (MMLV) vector (10, 11) and a lentiviral vector (pLV) (12, 13) in parallel to compare the pseudotyping efficiencies in these two related but different systems. The pLV vector pseudotyped both VSV-G (pLV-G) and SARS-CoV-2 spike glycoprotein (pLV-S) efficiently, whereas MMLV vector could only pseudotype VSV-G (Fig 1). The success of pseudotyping was assessed by the expression of green fluorescent protein (gfp) since both MMLV and pLV vectors integrate the gene encoding for gfp.

**Fig. 1.**
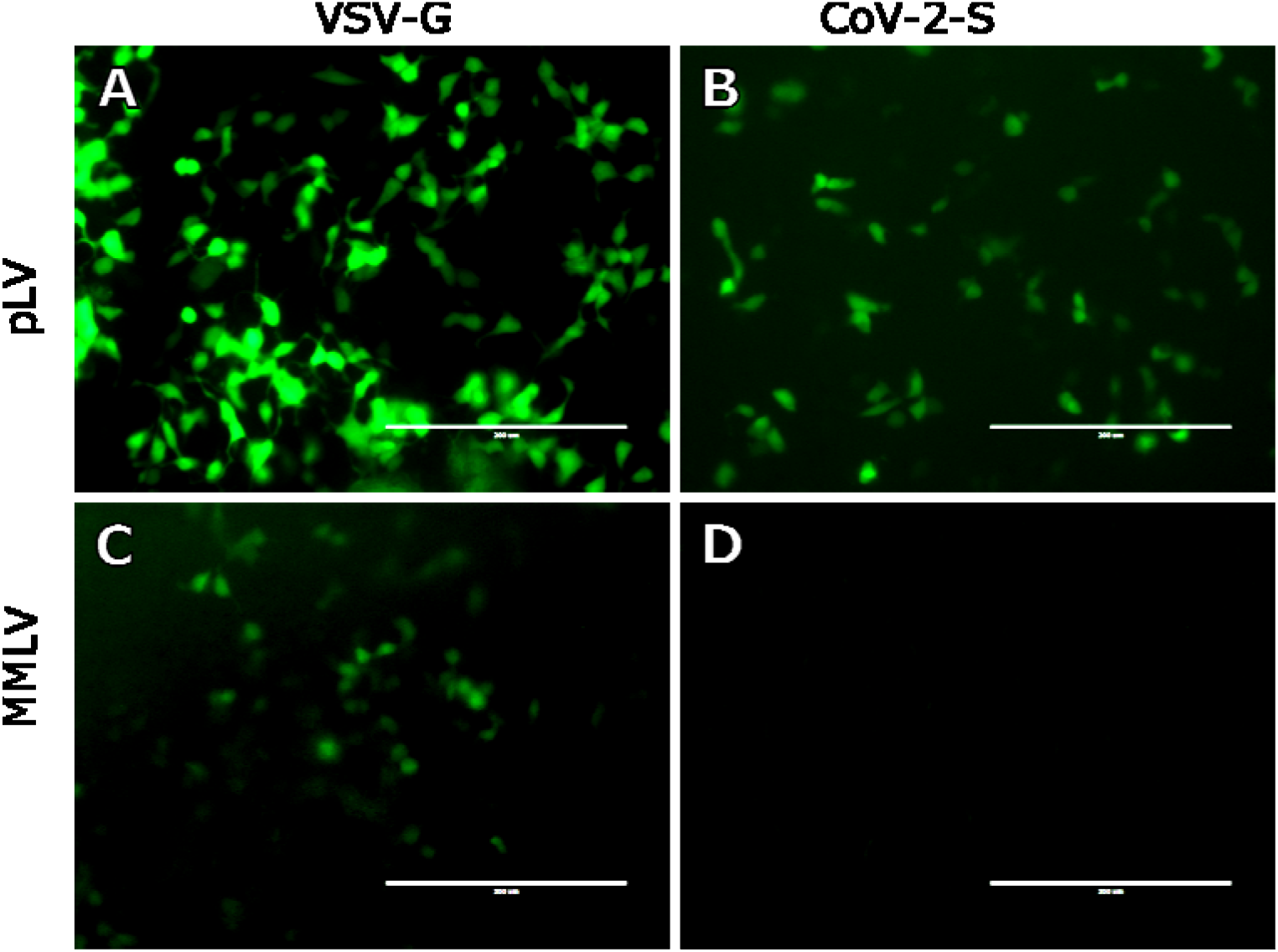
Transduction of HEK293T cells with lentiviral vector (pLV) pseudotyped with VSV-G (A) or CoV-2-S protein (B), and retroviral vector (MMLV) pseudotyped with VSV-G (C) or CoV-2-S (D). Both lentiviral and retroviral backbones incorporate enhanced green fluorescent protein (eGFP) that is expressed upon integration into target cells. The fluorescence was recorded at 48 hours post transduction. Magnification 20X. Scale bar:200 μm.

### The efficiency of SARS-CoV-2 SPG pseudotyped lentivirus transduction depends on cell type

We compared the efficiency of pLV-S transduction in 293T and VeroE6 cells since both of these cells are known to express the putative S receptor hACE2 on their surface in sufficient quantities to allow for virus entry (14). The 293T cells were transduced much more efficiently with pLV-G than pLV-S, whereas Vero E6 cells showed a low level of transduction for both viruses (Fig 2). We then analyzed the transduction efficiencies of adenocarcinomic human alveolar basal epithelial cells (A549), Madin-Darby Canine Kidney (MDCK), Vero E6 and 293T cells and compared them to 293T cells transfected with plasmid encoding hACE2 receptor. While untransfected 293T cells were half as efficient as hACE2 transfected cells, the MDCK, VeroE6 and A549 showed only a low level of transduction (Fig 2). Thus, the 293T cell line was used in the development of the neutralizing antibody assay.

**Fig. 2.**
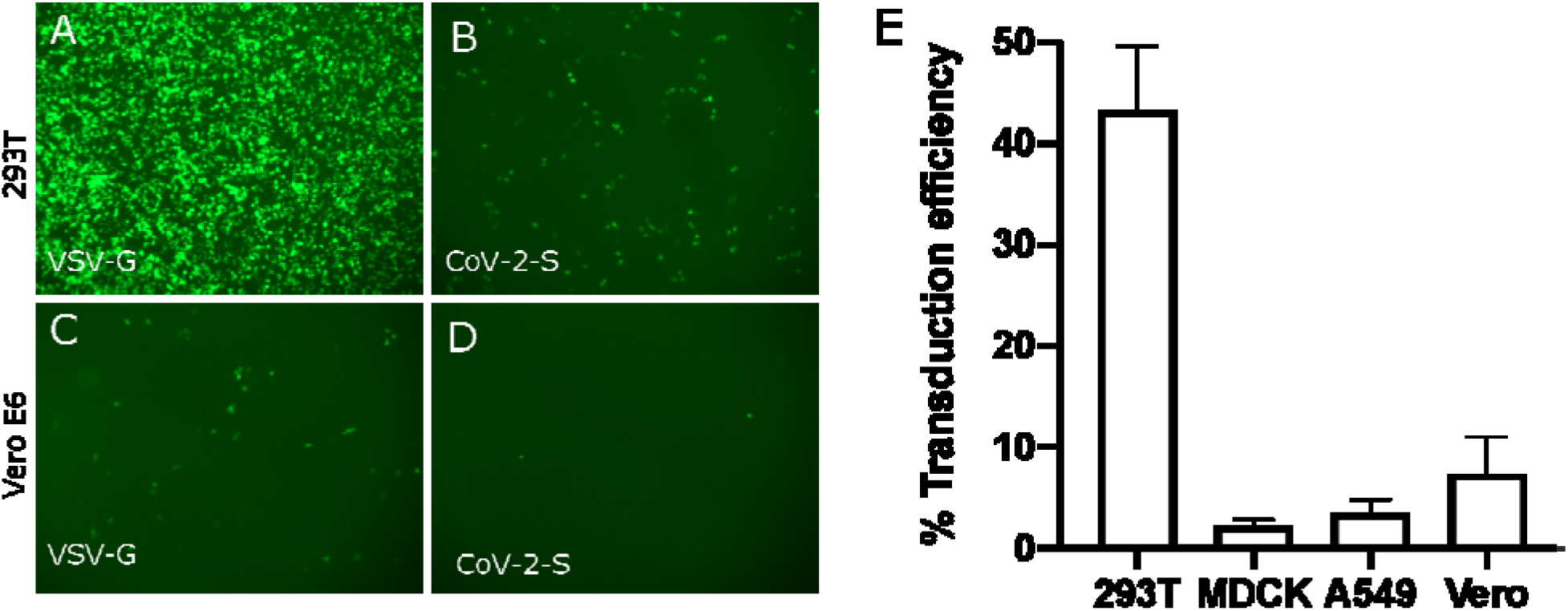
A) Transduction of pLV pseudotyped with VSV-G (A, C) or CoV-2 Spike glycoprotein (B, D) in HEK293T or Vero E6 cells. The lentiviral backbone incorporates enhanced green fluorescent protein (eGFP) that is expressed upon integration into target cells. The fluorescence was recorded at 48 hours post transduction. Magnification 4X. (E) Transduction efficiency of pLV pseudotyped with CoV-2 Spike glycoprotein in HEK293T, Madin-Darby Canine Kidney cells (MDCK), adenocarcinomic human alveolar basal epithelial cells (A549), and Vero (VeroE6) cells depicted as a percentage of efficiency in 293T cells transfected with plasmid encoding hACE2 receptor (hACE2-293T). The fluorescence was recorded at 48 hours post transduction. The experiments were done in triplicates and standard error of mean was plotted as error bars.

### Serum from COVID-19 convalescent patient effectively neutralizes pLV-S transduction

We tested the impact of patient serum on pLV-S transduction. Serial dilutions (1:40, 1:80, 1:160:1:320, 1:640 and 1:1280) of serum were made in the serum-free cell culture medium and incubated with equal amounts of pLV-S virus, then plated on 293T cells. Fluorescence images captured at 48 hours post infection showed significant reduction in pLV transduction with the serum from patient N304 but not from N308 (Fig. 3). Patient N304 tested positive in a RT-PCR assay for SARS-CoV-2, suffered a severe COVID-19 pneumonia and eventually recovered. Patient N308 suffered mild symptoms consistent with an upper respiratory infection but tested negative for SARS-CoV-2 infection by RT-PCR.

**Fig. 3.**
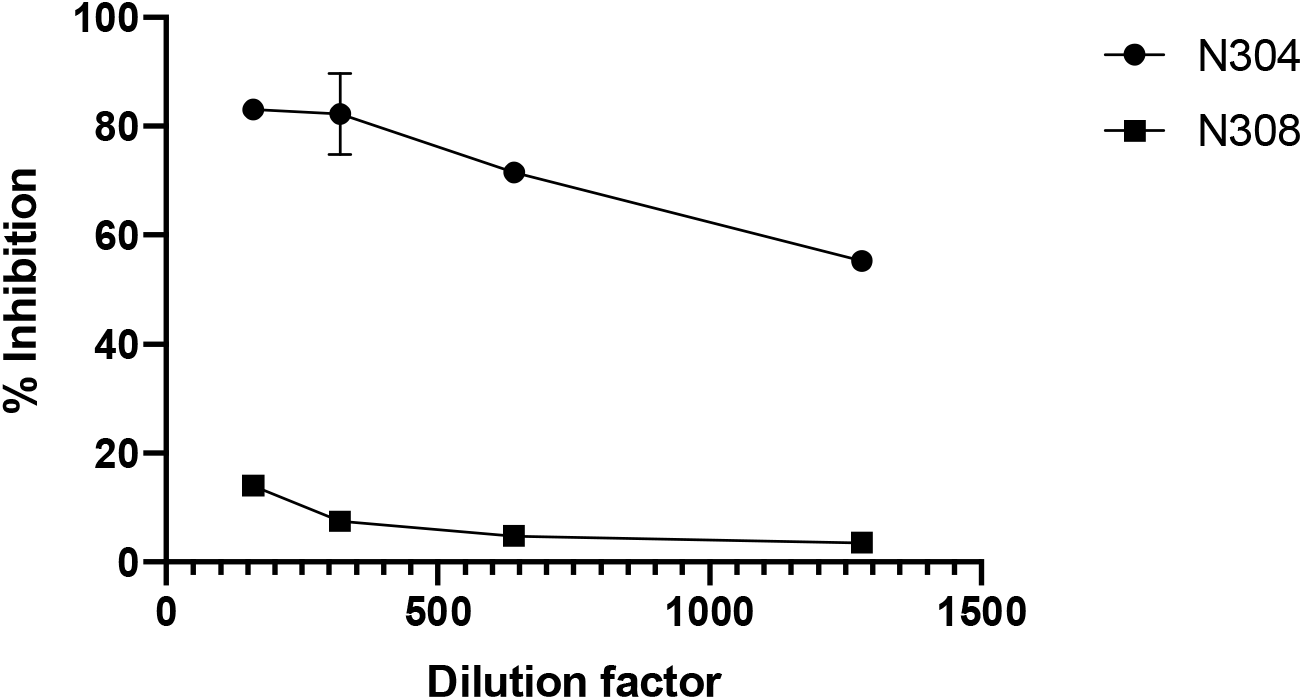
Neutralization of SARS-CoV-2 S glycoprotein pseudotyped pLV (pLV-S) using diluted patient serum. The serum was obtained from a convalescent patient (N304) or a mildly symptomatic individual (N308) at 30 days after onset of symptoms. Patient N304 tested positive for SARS-CoV-2 in a RT-PCR diagnostic test and N308 tested negative. Relative inhibition of pseudovirions at serial dilutions of patient serum compared to mock-serum control is shown. The fluorescence was recorded at 48 hours post transduction. The titers were performed in triplicates and standard error of mean was plotted as error bars.

## Discussion

Here we report the development of a lentiviral pseudotyping system for SARS-CoV-2 and its efficacy in detecting neutralizing antibody titers in convalescent patient serum. While a few pseudotyping systems for SARS-CoV-2 are in development, this report provides initial evidence that lentiviral pseudotyped SARS-CoV-2 can be used to determine neutralizing antibody titers in patient samples.

We first tried a traditional retroviral pseudotyping system based on MMLV. We have used this system earlier for creating cell lines stably expressing viral proteins of interest (15, 16). While the control VSV-G protein was pseudotyped successfully, we could not pseudotype SARS-CoV-2 SPG onto this backbone. Since retroviral vectors require actively dividing cells for successful transduction, we tested 293T cells at different levels of confluency but were unable to transduce them. The relative efficiency of VSV-G to transduce similar cells indicate that inherent qualities or complexity of SPG structure may make it more difficult to pseudotype it in this system.

Next, we utilized a third generation lentiviral system to pseudotype VSV-G and SPG. This system was able to pseudotype both glycoproteins although, as expected, efficiency was much better for VSV-G. The transduction efficiency would logically be guided by the level of SPG receptor expression on cells, therefore, we tested various mammalian cells lines that were available to us and are known express hACE2. We also transfected cells (293Ts) with hACE2 plasmid and compared transduction efficiency of different cells to these hACE2-293T. None of the cells transduced as efficiently as hACE2-293Ts and mock transfected 293Ts were only 42% efficient (Fig 2). Since the differences between hACE2-293Ts and 293Ts were only 2-2.4 fold, we decided to use 293Ts for patient serum screening.

Serum samples were obtained from two patients at University of Mississippi Medical Center. The first patient (N304) tested positive for SARS-CoV-2 in a RT-PCR assay and was convalescent at the time of sample collection. The second patient (N308) was mildly symptomatic and tested negative for SARS-CoV-2 (N308) in the same RT-PCR assay. Both serums were collected thirty days after onset of symptoms while patients were totally asymptomatic. Antibody testing in a SARS-CoV-2 SPG receptor binding domain ELISA revealed higher levels of RBD-specific IgM and RBD-specific IgG for N304 but lower levels for N308 (Table 1). Serial dilutions of serum N304 effectively neutralized pLV-S up to 1:1280 dilution whereas N308 did not neutralize the virus at higher than 1:80 dilution (Fig 3).

**Table 1.** The receptor binding domain (RBD) - specific IgM and IgG titers were measured in an ELISA using the recombinant RBD of the SARS-CoV-2 Spike protein as antigen.

| **Patient** | **IgM** | **IgG** |
| --- | --- | --- |
| N304 | 4,656 | 49,131 |
| N308 | 1,332 | 363 |

Thus, the pseudotyped virus (pLV-S) can be effectively used for screening of patient serum for the presence of neutralizing antibodies associated with acquired immunity to subsequent infection upon re-exposure. Determining functional immunity has potential social implications as the world community begins to recover from the tremendous social impact of the pandemic. Information regarding the development of herd immunity is important in this effort and can be gained from the results of this assay as well. These pseudoparticles can also be utilized for screening of inhibitors of SPG-hACE2 binding and resultant virus entry. They can also be utilized for screening of potential vaccine candidates as they represent SPG on their surface in its native confirmation.

## Methods

### Cells

293T (ATCC # CRL3216), Vero E6 (ATCC # CRL1586) and MDCK (ATCC #CCL-34) were cultured in DMEM (Corning Inc) supplemented with 10% fetal bovine serum (FBS, Fisher Scientific). A549 (ATCC #CCL-185) were cultured in F-12K (ATCC #30-2004) supplemented with 10% fetal bovine serum. 293T cells were transfected with hACE2 plasmid (a gift from Hyeryun Choe (Addgene plasmid # 1786; http://n2t.net/addgene:1786; RRID:Addgene_1786) to produce hACE2-293T cells. All mammalian cells were cultured at 37 °C with 5% CO_2_.

### ELISA

RBD-specific antibodies were measured generally according to the protocol developed in the Krammer Lab (17). Briefly, 384-well MaxiSorp plates (Thermo Fisher Scientific) were coated with purified recombinant RBD at concentration of three μg per ml in PBS. The coating volume and reaction volumes were 25 μl per well. Plates were incubated overnight at 4 °C, washed 3x with PBS-Tween20, and blocked with PBS containing 3% dry milk for one hour at room temperature. Blocking buffer was removed, and samples (one log dilutions in blocking buffer, 5×10^1^-5×10^4^) were added to the wells. Plates were incubated for two hours at room temperature before washing three times. Horse radish peroxidase conjugated to anti-human IgG FC (Southern Biotech) or anti-human IgM (Southern Biotech) was diluted 1:2,000 in PBS containing 1% dry milk, added to the wells, and incubated at room temperature for one hour. Plates were washed five times and developed with tetramethyl-benzidine (Southern Biotech). After 30 minutes, development was stopped by adding 25 μl of 2N H_2_SO_4_ to each well. Absorbance was measured at 450 nm. The end point dilution titer is the serum dilution that results in an absorbance of 0.2 absorbance units over background.

### Generation of pseudotype particles

HEK293T cells (2 x 10^6^) were plated in a 100-mm tissue culture dish and transfected the next day when they were about 75% confluent with a combination of the following plasmids: 9 μg of *pLV-eGFP* (a gift from Pantelis Tsoulfas (Addgene plasmid # 36083; http://n2t.net/addgene:36083; RRID:Addgene_36083), 9 μg of *psPAX2* (a gift from Didier Trono (Addgene plasmid # 12260; http://n2t.net/addgene:12260; RRID:Addgene_12260), and 3 μg of *pCAGGS-S (SARS-CoV-2)*(Catalog No. NR-52310:BEI Resources) or VSV-G (a gift from Tannishtha Reya (Addgene plasmid # 14888; http://n2t.net/addgene:14888; RRID:Addgene_14888) as control. Polyethylenimine (PEI) reagent (Millipore Sigma, #408727) was used for transfection following manufacturer’s protocols. Next day, the cells were checked for transfection efficiency under a fluorescent microscope, indicated by GFP fluorescence. The supernatants from cell culture at 24 h were harvested and stored at 4 °C and more (10 ml) complete media (DMEM + 10% FBS) was added to the plates. The supernatant from cell culture at 48 h was harvested and combined with the 24 h supernatant for each sample. The combined supernatants were spun in a tabletop centrifuge for 5 min at 2000 g to pellet the residual cells and then passed through a 0.45 micron syringe filter. Aliquots were frozen at −80 °C. New 293T cells plated in 12 well tissue culture dish were infected with the harvested virus (supernatant) with a dilution range of 10^2^ to 10^7^. Virus titers were calculated by counting the GFP positive cells in the dilution with 20-100 GFP positive cells. MMLV pseudovirions were made in a similar way. HEK293T cells were transfected with the following plasmids:10 μg pLNCX-GFP, 1.2 μg pCMV-tat-HIV, 6 μg pJK3, and 3 μg of pL-VSV-G (control) or pCAGGS-S (SARS-CoV-2). Supernatants were recovered in a fashion similar to above.

### Patient serum screening

Samples were obtained either from discarded clinical samples or from individual recruited to participate in this study. Written informed consent was obtained from the individuals recruited for this study. The study was reviewed and approved by the Institutional Review Board of the University of Mississippi Medical Center which follows the national and international guidelines consistent with the principles established by the Declaration of Helsinki. Serial dilutions of the patient serum samples (1:40; 80; 160; 320; 640 and 1280) were made in DMEM with end volume of 60 μl each. Sixty μl of the supernatant stock (diluted to give 200-500 GFP + cells/well) was mixed with the serum and incubated for 1 h at 37 °C. Stock + serum dilutions were laid over 293T cells plated in 96 well tissue culture dishes and incubated at 37 °C and 5% CO_2_ for 2 h. Medium was replaced with complete medium (DMEM + 10% FBS) and incubated for another 48 h. The assay was read and analyzed using Lionheart FX automated fluorescent microscope (BioTek Instruments, Inc., Winooski, VT, USA).

## Data Availability

Non identifiable data is included in the manuscript.

## Acknowledgments

We would like to acknowledge the receipt of pCAGG-S and pCAGGS-RBD plasmids from Florian Krammer at Icahn School of Medicine at Mount Sinai Hospital, NY. Funding for this work was provided by University of Mississippi Medical Center COVID-19 funds.

